# Effects of Cannabidiol on Auditory Mismatch Negativity in Autistic Children

**DOI:** 10.64898/2026.10.14.26365371

**Authors:** Karan Budhiraja, Nina-Francesca Parrella, Aron T. Hill, Luke A. Downey, Sarah Catchlove, Peter G. Enticott, Talitha C. Ford

**Affiliations:** Cognitive Neuroscience Unit, School of Psychology, Deakin University, Burwood, Australia; Centre for Mental Health & Brain Sciences, School of Health Sciences, Swinburne University of Technology, Melbourne, Australia; Eastern Health Clinical School, Monash University, Melbourne, Australia

## Abstract

Pharmacological treatments targeting core autistic features remain limited. In neurotypical adults, cannabidiol (CBD) alters mismatch negativity (MMN), an electrophysiological marker of automatic auditory deviance detection that has been associated with autistic traits. The effects of CBD on MMN, as measured using electroencephalography (EEG), in autistic children remain unknown. This secondary analysis of a pilot randomised, double-blind, placebo-controlled crossover trial examined the effects of 12 weeks of oral CBD oil on MMN during a duration-deviant auditory oddball task. We hypothesised that CBD would produce more negative MMN amplitudes and shorter peak latencies relative to placebo. Nineteen autistic children (6-12 years, 8 female) received CBD (dose) and placebo in randomised order, separated by an 8-week washout. Age-adjusted linear mixed-effects models tested Time (baseline, follow-up) × Treatment (CBD, placebo) effects on mean MMN amplitude and peak latency separately at Fz and FCz. Exploratory Spearman correlations examined baseline associations between MMN and Developmental Behaviour Checklist-2 (DBC-2) Social Relating scores. At FCz, latency decreased following CBD but increased following placebo but did not survive FDR correction (*p* = .029*, p_FDR_* = .058) and no other interactions were significant. Higher baseline DBC-2 Social Relating scores were associated with more negative MMN amplitude at FCz (*ρ* = -.58, *p*_FDR_ = .028) and Fz (*ρ* = -.59, *p_FDR_* = .028). These findings provide tentative evidence that CBD may modulate MMN latency in autistic children. Larger trials should clarify CBD’s effects on MMN and further examine the association between MMN amplitude and social-relating difficulties.

## Introduction

Automatic detection of changes in the auditory environment supports adaptive perception and effective communication (Poulsen et al., 2024). In autism, auditory processing differences have been repeatedly associated with speech, language, and broader social communication challenges (Baranek et al., 2019; Key & D’Ambrose Slaboch, 2021; Poulsen et al., 2024). Yet pharmacological interventions have largely targeted co-occurring behavioural and psychiatric difficulties, including impulsivity, anxiety, and depression (Baldes et al., 2026; Manter et al., 2025), leaving auditory processing differences relatively unaddressed. Cannabidiol (CBD), a non-intoxicating compound derived from the *Cannabis sativa* plant, has emerged as a potential intervention in autism, with preliminary studies reporting improvements in social functioning, disruptive behaviour, anxiety, and parental stress following multi-week administration (Aran et al., 2021; Parrella et al., 2026; Silva Junior et al., 2024). The use of caregiver- or clinician-reported measures is susceptible to expectancy effects (Jones et al., 2017), leaving it unclear whether such changes are accompanied by altered neural function. This study examines whether 12-weeks of CBD oil can modulate the mismatch negativity (MMN), an objective electroencephalography (EEG)-derived event-related potential (ERP) that indexes automatic auditory deviance detection and has frequently been reported as atypical in autistic children (Näätänen et al., 2007; Sapey-Triomphe et al., 2025; Schwartz et al., 2018). By linking CBD to an objective neural measure, this study assesses whether the reported behavioural changes are accompanied by measurable alterations in auditory neurophysiology and evaluates MMN as a potential marker of treatment-related neural change.

Sensory processing differences are recognised in the diagnostic criteria for autism and occur across auditory, visual, tactile, olfactory, and proprioceptive modalities (American Psychiatric Association, 2013; Kadlaskar et al., 2023). Reported in an estimated 45-95% of autistic individuals, these differences can alter how sensory information is detected, interpreted, and responded to (Ben-Sasson et al., 2019; Thye et al., 2018). Early sensory differences are associated with social functioning in autism and predict later difficulties in socialisation, communication, and adaptive functioning (Grzadzinski et al., 2020; Williams et al., 2018; Worthley et al., 2024). Such differences are also associated with emotion regulation difficulties, maladaptive behaviours, and poorer mental health outcomes (Chen et al., 2025; Rossow et al., 2023). These findings suggest that sensory processing differences may be an early-emerging factor associated with broader social, behavioural, and emotional functioning across development in autism.

Among these sensory domains, auditory processing differences are particularly relevant to social communication (Poulsen et al., 2024; Thye et al., 2018). Effective social communication depends on auditory processing at multiple levels, from orienting to socially relevant sounds and discriminating acoustic changes to interpreting higher-order cues such as vocal prosody that convey emotion and communicative intent (Belyk & Brown, 2014; Hellbernd & Sammler, 2018). Autistic individuals show differences across these levels, including reduced spontaneous orienting to speech (Key & D’Ambrose Slaboch, 2021), atypical automatic detection of speech-sound changes (Chen et al., 2020; Ludlow et al., 2014; Sapey-Triomphe et al., 2025), and poorer recognition of emotion from vocal cues (Leipold et al., 2023; Luo et al., 2026). Importantly, early-stage auditory processing has been associated with language and communication outcomes in autistic children (Cacciato-Salcedo et al., 2026; Key & D’Ambrose Slaboch, 2021), suggesting that some higher-order social communication difficulties may partly reflect differences arising at earlier stages of auditory analysis.

Pre-attentive deviance detection is a key early-stage auditory process indexed by the MMN. It is typically elicited using oddball paradigms, in which infrequent deviant stimuli violate regularities established by repeated standards, largely independently of directed attention (Chen et al., 2020; Näätänen et al., 2007). Because it requires no overt behavioural response, the MMN is well suited to assessing auditory processing in children and neurodevelopmental populations with varying verbal and cognitive abilities (Csépe & Honbolygó, 2024; Parrella et al., 2023). Studies in autistic children have yielded mixed results, including larger, smaller, and comparable MMN amplitudes and shorter, longer, and comparable MMN latencies relative to typically developing peers (e.g., Kabil et al., 2023; Lassen et al., 2022; Matsuba et al., 2024; Mayerle et al., 2023). Nevertheless, recent reviews indicate generally attenuated MMN amplitudes and prolonged latencies in autistic children, particularly in non-speech paradigms involving frequency and duration deviants, with such pattern less consistent in adults (Cacciato-Salcedo et al., 2026; Chen et al., 2020; Sapey-Triomphe et al., 2025). Despite the age- and paradigm-related differences, MMN may still be useful for assessing within-individual neural changes, as illustrated by pharmacological studies in schizophrenia (O’Donnell et al., 2023; Sehatpour et al., 2022).

Predictive processing provides a framework for understanding MMN variability in autism (Cacciato-Salcedo et al., 2026; Sapey-Triomphe., 2025). It proposes that the brain maintains a hierarchical generative model of its environment, predicting incoming sensory input and minimising prediction errors that arise when the input deviates from these predictions (Auksztulewicz & Friston, 2015). Accordingly, the MMN is conceptualised as a precision-weighted prediction error, with precision (i.e., estimated reliability) regulating the influence of sensory evidence relative to prior expectations (Auksztulewicz & Friston, 2015; Fitzgerald & Todd, 2020). Autism has been proposed to involve atypical precision weighting (Coll et al., 2020; Van De Cruys et al., 2014), with some findings suggesting reduced influence of prior expectations on sensory processing (Chouinard et al., 2018; Wagley et al., 2020). However, broader research indicates that autistic children can use learned regularities to anticipate upcoming sensory input, while evidence for overweighted sensory prediction errors remains inconsistent (Papastamou et al., 2025; Prescott et al., 2024; Van De Cruys et al., 2021). Recent reviews suggest that autism may involve context-dependent differences in precision weighting rather than a uniform directional imbalance (Angeletos Chrysaitis & Seriès, 2023; Cannon et al., 2021; Cui et al., 2026). Such differences may contribute to MMN variability, raising questions about the neural mechanisms underlying precision weighting and whether they can be modified.

Neurobiologically, precision weighting is proposed to be implemented through modulation of postsynaptic gain, or responsiveness, of superficial pyramidal cells encoding prediction errors, such that greater gain increases the influence of sensory evidence on perceptual updating (Brown & Friston, 2012; Lawson et al., 2014; Sapey-Triomphe et al., 2021). This gain is partly regulated by excitatory-inhibitory (E/I) mechanisms, including N-methyl-D-aspartate receptor (NMDAR)-dependent engagement of gamma-aminobutyric acid (GABA) inhibitory circuitry (Ferguson & Cardin, 2020; Lewis et al., 2022). In autism, GABAergic alterations have been observed across multiple levels of cortical organisation including receptor expression (Hong et al., 2020), signalling in the visual cortex (Robertson et al., 2016), and GABA and glutamate neurometabolite concentrations in the auditory cortex (Du et al., 2023; Port et al., 2017). Growing evidence links altered GABAergic regulation and its potential effects on cortical gain to sensory and communication differences in autism (Ford et al., 2017, 2020; Johnson et al., 2023; Kirkovski et al., 2018; Orekhova et al., 2019; Puts et al., 2017; Yizhar et al., 2011). Beyond these associations, pharmacological enhancement of GABAergic inhibition has been shown to modulate sensory cortical gain (Pagano et al., 2023; Schallmo et al., 2018), raising the possibility that E/I mechanisms represent a shared therapeutic target for sensory and communication difficulties in autism.

Preclinical evidence suggests that pharmacological modulation of GABAergic signalling can ameliorate autism-related behavioural phenotypes (Han et al., 2014; Silverman et al., 2015). The endocannabinoid system (ECS) helps regulate the balance between GABAergic and glutamatergic signalling (Scheyer et al., 2023). Through activity-dependent retrograde signalling, endocannabinoids such as anandamide (AEA) act on presynaptic CB1 receptors to modulate GABA and glutamate release, thereby influencing E/I dynamics and cortical responsiveness (Zou & Kumar, 2018). ECS signalling may also be altered in autism, as suggested by lower circulating AEA concentrations in autistic individuals (Jia et al., 2025). Experimentally increasing endocannabinoid levels has improved behavioural outcomes in animal models, while pharmacological inhibition of AEA degradation has increased circulating AEA in autistic adolescents and adults, with greater increases associated with improved social responsiveness (Jia et al., 2025; Klein et al., 2025). Collectively, these findings support the investigation of the ECS as a target for pharmacological modulation.

CBD can modulate the ECS and other molecular targets implicated in E/I regulation (Manzoni et al., 2025). For example, CBD altered glutamatergic and GABAergic neurochemistry in autistic and non-autistic adults following acute administration (Pretzsch et al., 2019) and potentiated GABA-mediated currents in human cortical tissue in vitro (Ruffolo et al., 2022). Preclinical studies similarly link CBD-induced enhancement of GABAergic signalling to improvements in autism-relevant social and behavioural features (Kaplan et al., 2017; Zhang et al., 2023). Evidence further suggests that CBD-based interventions may improve selected social and behavioural outcomes in autistic children, although robust evidence remains limited (Aran et al., 2021; Hacohen et al., 2022; Parrella et al., 2026; Silva Junior et al., 2024). Importantly, CBD has also been shown to modulate the MMN, with acute administration increasing its amplitude in response to duration and intensity deviants in adults with infrequent cannabis use (Greenwood et al., 2022). However, whether CBD affects MMN in autistic children remains unknown.

Accordingly, we aimed to determine whether 12 weeks of CBD administration modulated MMN amplitude and latency in autistic children. Building on this rationale and previous findings of generally attenuated MMN amplitude and prolonged latency in this population (Cacciato-Salcedo et al., 2026; Sapey-Triomphe et al., 2025), we hypothesised that the CBD intervention would produce more negative MMN amplitudes and shorter latencies relative to placebo. We also explored whether baseline MMN amplitude and latency were associated with parent-reported social-relating scores.

## Materials and methods

This study was a secondary analysis of electroencephalography (EEG) and behavioural data from a larger randomised, double-blind, placebo-controlled crossover trial registered with the Australian New Zealand Clinical Trials Registry (ACTRN12622000437763). Participants completed two 12-week intervention periods, receiving CBD and placebo in a randomised order, separated by an 8-week washout period (see Figure 1). EEG recordings and parent-reported behavioural measures were obtained before and after each intervention period. Further details are reported in the original trial paper (Parrella et al., 2026). Ethics approval was provided by the Deakin University Human Research Ethics Committee (DUHREC 2020-071). Written informed consent was obtained from parents or guardians, along with assent from participants.

**Figure 1.**
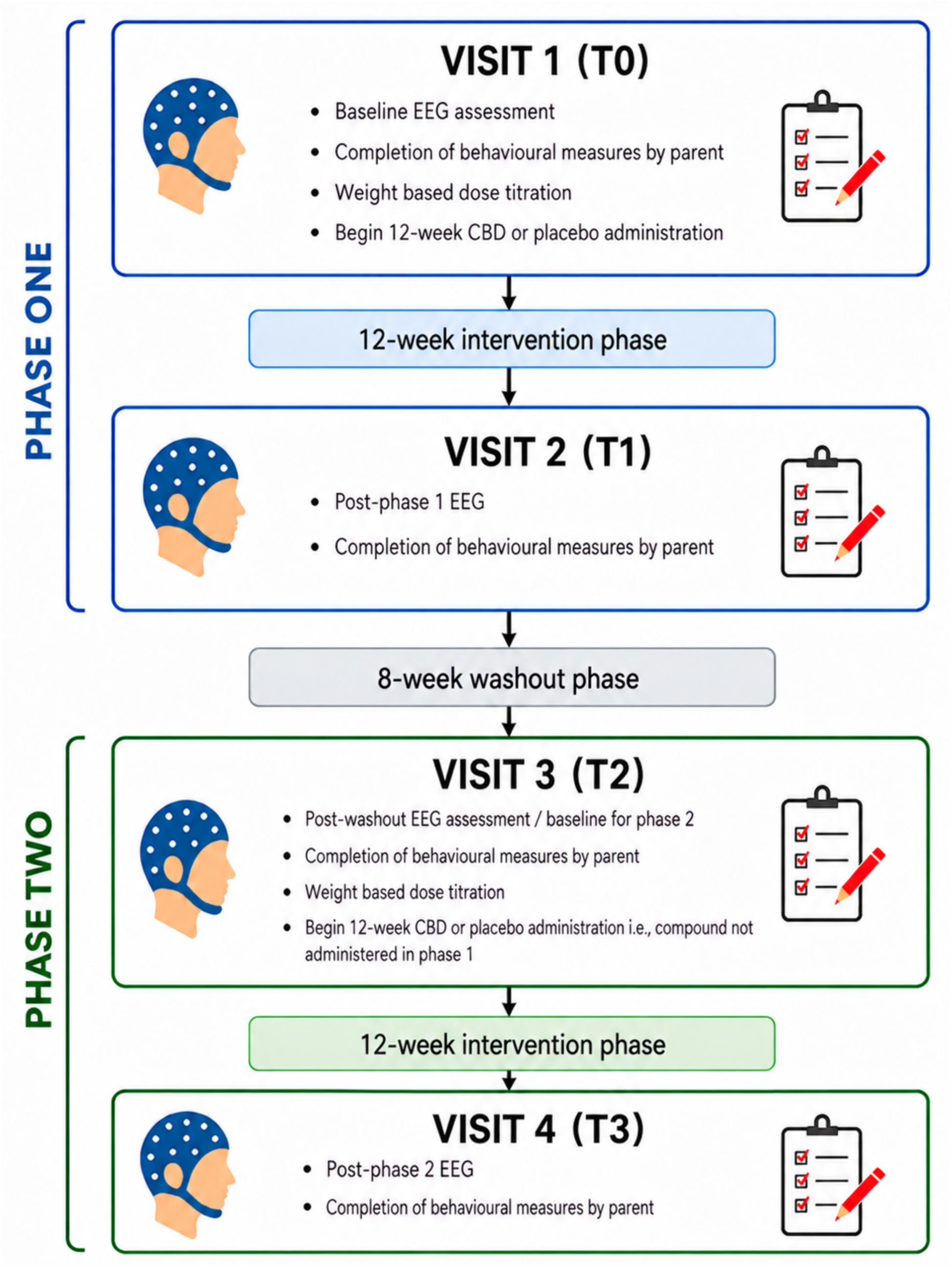
Study Design and Assessment Timeline. CBD = cannabidiol; EEG = electroencephalography; T0 = initial baseline; T1 = phase 1 post-intervention; T2 = phase 2 baseline (post-washout); T3 = phase 2 post-intervention. Adapted from Parrella et al. (2026).

## Participants

Of the 34 autistic children enrolled in the larger trial, 20 underwent EEG recording (aged 6-12, 8 female). Prior to enrolment, autism diagnoses were confirmed by the participants’ diagnosing clinicians using standardised assessment instruments (e.g., the Autism Diagnostic Observation Schedule, parent interview) and review of medical records. Participants continued their usual medications and behavioural therapies throughout the study.

## Intervention

The intervention was administered as a broad-spectrum CBD oil (Medigrowth CBD100), containing 100 mg/mL cannabidiol and naturally occurring cannabis-derived terpenes, with negligible delta-8-tetrahydrocannabinol (THC) concentrations (<.01%). Dosage commenced at 5 mg/kg/day and was gradually titrated to a maximum of 10 mg/kg/day, with doses administered orally by trained caregivers. The placebo was supplied in identical bottles and matched the CBD oil in appearance and taste. Caregivers completed daily treatment logs during each intervention period to monitor treatment adherence and adverse effects. Further details are reported in Parrella et al. (2026).

## Experimental Procedure

During EEG recording, participants completed a passive auditory oddball paradigm to elicit the MMN. The paradigm comprised three blocks of 400 trials, each containing 340 standard tones (85%) and 60 deviant tones (15%). Standard tones were 50 ms in duration, whereas deviant tones were 100 ms. Both were presented at 1000 Hz and 70 dB, with a 500 ms interstimulus interval. Trials were pseudorandomised such that at least two standards occurred between successive deviants. Auditory stimuli were delivered using E-Prime (v3.00) via the computer’s built-in speakers. Speakers were used rather than head/earphones to improve tolerability and accessibility for participants. During the task, participants were instructed to quietly engage with the muted Toca Kitchen iPad application while ignoring the auditory stimuli. This concurrent activity was intended to support engagement and tolerability during EEG acquisition. The paradigm lasted approximately 15 minutes.

## EEG Recording and Processing

### EEG Acquisition

EEG was recorded using a 64-channel HydroCel Geodesic Sensor Net (Electrical Geodesics Inc., Eugene, OR, USA) and a NetAmps 4000 amplifier at a sampling rate of 1000 Hz. Signals were referenced online to the vertex (Cz). The net used Ag/AgCl electrodes embedded in electrolyte-soaked sponges. Impedances were maintained below 50 kΩ during EEG recordings. During the MMN paradigm, participants were seated in a dimly lit room while trained research staff monitored them throughout the task to minimise movement.

### EEG Preprocessing

EEG data were preprocessed using custom scripts in MATLAB (R2024b; The MathWorks, Inc., 2024), incorporating EEGLAB (v2026.0.0; Delorme & Makeig, 2004) and ERPLAB (v13.00; Lopez-Calderon & Luck, 2014). Channels were relabelled according to their approximate equivalents in the international 10-10 system, and four peripheral electrodes (E61-E64) were removed, leaving 60 scalp channels. Artefacts were attenuated using RELAX-Jr (Hill et al., 2024), an automated preprocessing pipeline adapted from RELAX (Bailey et al., 2023) for paediatric EEG data acquired using Geodesic Sensor Net caps. Multichannel Wiener filtering attenuated muscle, blink, horizontal eye-movement, and drift artefacts (Somers et al., 2018). Independent component analysis (ICA) was then computed using the PICARD algorithm (Ablin et al., 2018), and components identified using the adjusted-ADJUST classifier (Leach et al., 2020) were cleaned using wavelet-enhanced ICA (wICA; Castellanos & Makarov, 2006). Any bad electrodes removed during cleaning were then interpolated back into the record using spherical interpolation.

Following automated preprocessing, we visually inspected continuous EEG traces and power spectra for residual artefacts and overall data quality. All recordings from one participant were removed due to excessively noisy EEG traces for each timepoint. Of the remaining 19 participants (8 female), two recordings were removed as they lacked usable photosensor triggers, preventing reliable time-locking to stimulus onset (see Supplementary material). ERPLAB Binlister was used to classify auditory events as duration-deviant trials or standard trials immediately preceding each deviant. Events were time-locked to photosensor triggers marking stimulus onset. Data were subsequently segmented using RELAX-Jr into epochs from -100 to 400 ms relative to stimulus onset. Each epoch was baseline corrected by subtracting the mean voltage during the pre-stimulus interval from -100 to 0 ms. Epochs were rejected if absolute voltages exceeded 100 μV, or if improbable data or kurtosis values exceeded the threshold of 5 SD for any single channel, or 3 SD across all channels (Delorme & Makeig, 2004).

For each recording, retained epochs were averaged separately for the standard and duration-deviant conditions to generate ERPs. MMN difference waveforms were calculated by subtracting the standard ERP from the duration-deviant ERP. We visually inspected the resulting waveforms for data quality and excluded nine additional recordings because of excessive noise or insufficient retained trial counts (see Supplementary material).

### MMN Quantification

Grand-average standard and duration-deviant ERPs were generated across retained recordings, and the grand-average MMN difference waveform was calculated as deviant minus standard (see Figure 2). Based on the established frontocentral distribution of duration MMN and previous studies examining MMN at Fz and FCz (Le Sommer et al., 2023; Lee et al., 2020), the grand average was examined at these sites within a 100-250 ms interval. The most negative peaks were identified at 158 and 166 ms at Fz and FCz, respectively. Accordingly, 100-ms measurement windows centred on these peaks were defined as 108-208 ms at Fz and 116-216 ms at FCz (see Figure 2). Within each window, mean MMN amplitude was quantified as the average difference-wave voltage, while peak latency was defined as the latency of the most negative peak. Both measures were extracted separately at Fz and FCz for each retained recording.

**Figure 2.**
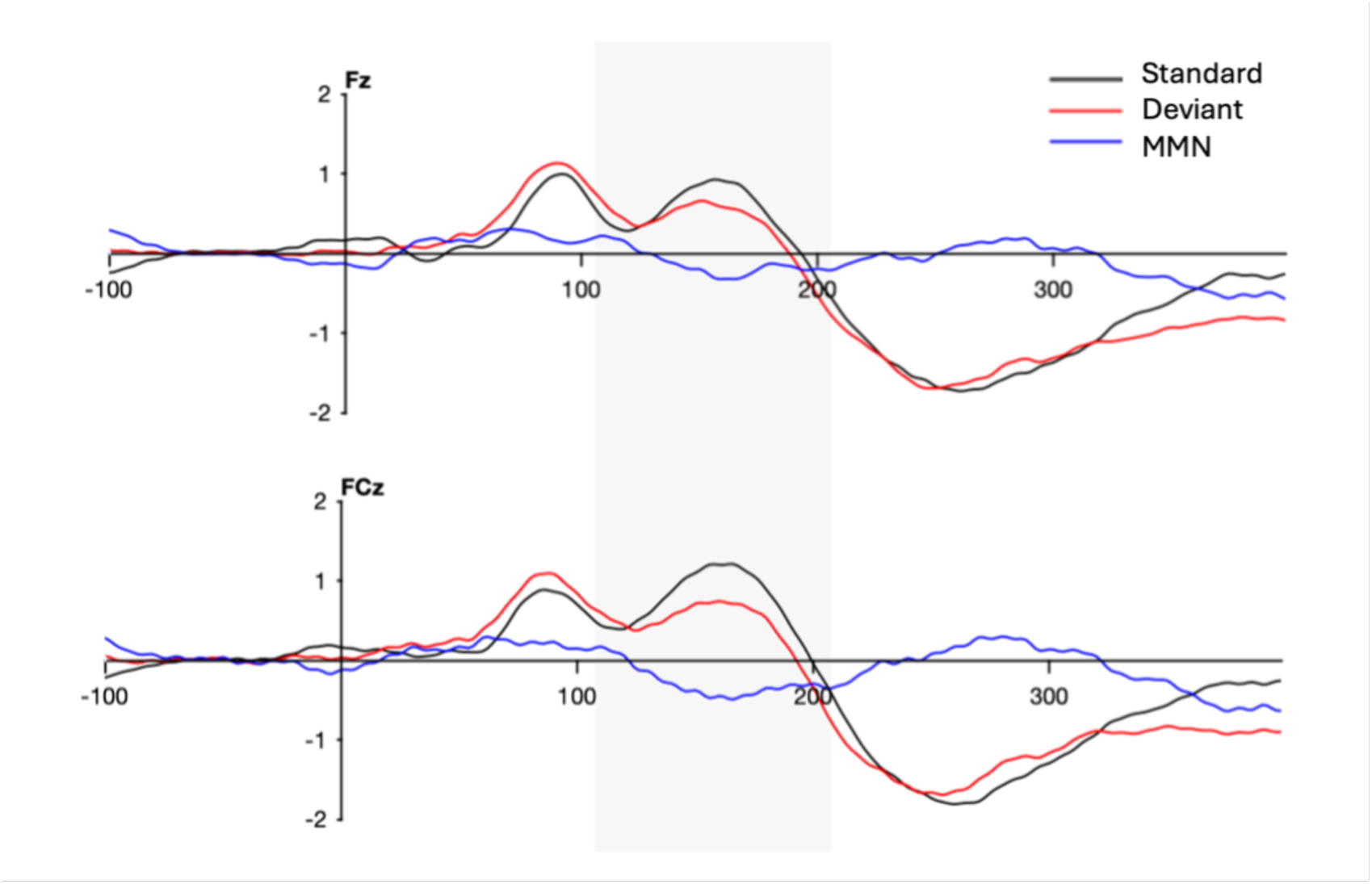
Grand-average standard, duration-deviant, and MMN waveforms at Fz and FCz. MMN was calculated as duration deviant minus standard. Time 0 ms represents stimulus onset. Negative voltage is plotted downward. Shading represents 100 ms measurement window of the mismatch negativity (MMN): Fz = 108-208 ms; FCz 116-216 ms.

## Developmental Behaviour Checklist-Second Edition

The Developmental Behaviour Checklist-Second Edition (DBC-2; Einfeld & Tonge, 2002) is a 96-item parent-report measure of behavioural and emotional difficulties in children and adolescents with developmental disabilities. Items are rated on a three-point scale from 0 (*not true as far as you know)* to 2 *(often true or very true)* and yield a total score and five subscale scores. We used raw scores from the 10-item Social Relating subscale (range = 0-20), with higher scores indicating greater social-relating difficulties. Exploratory correlational analyses examined associations between baseline Social Relating scores (T0; see Figure 1) and MMN measures.

## Statistical analysis

Data were analysed using jamovi (Version 2.7.37.0), with statistical significance set at *α* = .05. Linear mixed-effects models were fitted to all available EEG observations after quality-control exclusions (see Supplementary material). These models accounted for repeated observations within participants across treatment conditions and assessment time points and accommodated incomplete repeated-measures data under the missing-at-random (MAR) assumption. Separate models were fitted for MMN mean amplitude and peak latency at Fz and FCz. Each model included Time (baseline, 12 weeks), Treatment (CBD, placebo), and the Time × Treatment interaction as fixed effects, with age included as a covariate to account for developmental variation in MMN responses (Bishop et al., 2011; Themas et al., 2023). Participant was included as a random intercept to account for within-participant dependence. The Time × Treatment interaction was the effect of interest and tested whether change in MMN from baseline to 12 weeks differed between the CBD and placebo groups.

An exploratory follow-up model of MMN peak latency additionally included Electrode (Fz, FCz) and its interactions with Time and Treatment. The Time × Treatment × Electrode interaction tested whether treatment-related change differed between electrodes. Residual normality was assessed using Shapiro-Wilk tests and Q-Q plots. Although Shapiro-Wilk tests indicated departures from normality for the Fz amplitude (*p* = .017) and latency (*p* = .012) models, the Q-Q plots showed broadly acceptable distributions, with deviations primarily in the tails. Given the robustness of linear mixed-effects models to modest non-normality (Schielzeth et al., 2020), outcomes were analysed without transformation.

Using T0 baseline data (see Figure 1), exploratory age-adjusted partial Spearman correlations examined associations between DBC-2 Social Relating scores and MMN mean amplitude and peak latency at Fz and FCz. We used a rank-based approach because of its reduced sensitivity to non-normality and outliers, particularly given the small sample and variability typical of paediatric autism EEG data (De Winter et al., 2016; Webb et al., 2015). To account for multiple testing, we applied the Benjamini–Hochberg false discovery rate (FDR) correction separately to four families of tests comprising Time × Treatment effects for MMN amplitude, Time × Treatment effects for MMN latency, baseline associations with MMN amplitude, and baseline associations with MMN latency. FDR-adjusted *p* values below .05 were considered statistically significant.

Because this study was a secondary analysis of the parent trial, no separate *a priori* sample size calculation was conducted for the MMN outcomes. We therefore conducted a supplementary simulation-based sensitivity analysis using the observed sample of 19 participants and 59 EEG recordings (see Supplementary material). Simulations were conducted in R (Version 4.6.1; R Core Team, 2026) using the lme4 (Bates et al., 2015) and lmerTest (Kuznetsova et al., 2017) packages. The analysis estimated the interaction magnitudes corresponding to 80% power after FDR correction at .05, expressed as the between-group difference in change from baseline to 12 weeks in the original outcome units (ms or μV). Simulations retained the observed recording pattern and incorporated the fitted model parameters described above. For each electrode-specific estimate, the Time × Treatment interaction at the other electrode was set to zero while retaining the same FDR correction procedure. Approximate interaction magnitudes corresponding to 80% power were 44 ms for FCz latency, 48 ms for Fz latency, 1.3 μV for FCz amplitude, and 1.4 μV for Fz amplitude. These estimates were conditional on the fitted variance parameters and simulation assumptions.

## Results

### Participant characteristics

Of the 19 participants included in the EEG analyses, 10 contributed recordings at all timepoints, while the remaining nine contributed data from at least one timepoint (see Supplementary material). Table 1 presents the demographic and clinical characteristics of the sample. Four participants (21.1%) had a co-occurring intellectual disability. Baseline parent-reported Social Relating scores varied considerably across the sample.

**Table 1.** Participant Characteristics and Baseline DBC-2 Social Relating Scores.

| Characteristic | Category | *N* (%) | Mean (*SD*), range |
| --- | --- | --- | --- |
| Age, years | Total | 19 (100) | 9.59 (1.87), 6-12 |
|  | Male | 11 (57.9) | 9.53 (2.05), 6-12 |
|  | Female | 8 (42.1) | 9.67 (1.72), 7-11 |
| Body weight, kg | Total | 19 (100) | 37.89 (15.67), 26-62 |
|  | Male | 11 (57.9) | 36.45 (16.75), 26-61 |
|  | Female | 8 (42.1) | 39.88 (14.93), 21-62 |
| Intellectual disability diagnosis | Total | 4 (21.1) |  |
|  | Male | 1 (25.0) |  |
|  | Female | 3 (75) |  |
| DBC-2 Social Relating (T0) | Total | 15 (100) | 9.20 (4.81), 1-17 |
|  | Male | 7 (46.7) | 8.00 (4.44), 3-14 |
|  | Female | 8 (53.3) | 10.25 (5.18), 1-17 |

*Note.* Percentages are calculated using the number of participants with available data for each characteristic. DBC-2 = Developmental Behaviour Checklist-Second Edition; T0 = initial baseline assessment.

## MMN analyses

Table 2 summarises estimated marginal means and Time × Treatment effects for MMN amplitude and latency at Fz and FCz, with corresponding plots shown in Figure 3.

**Figure 3.**
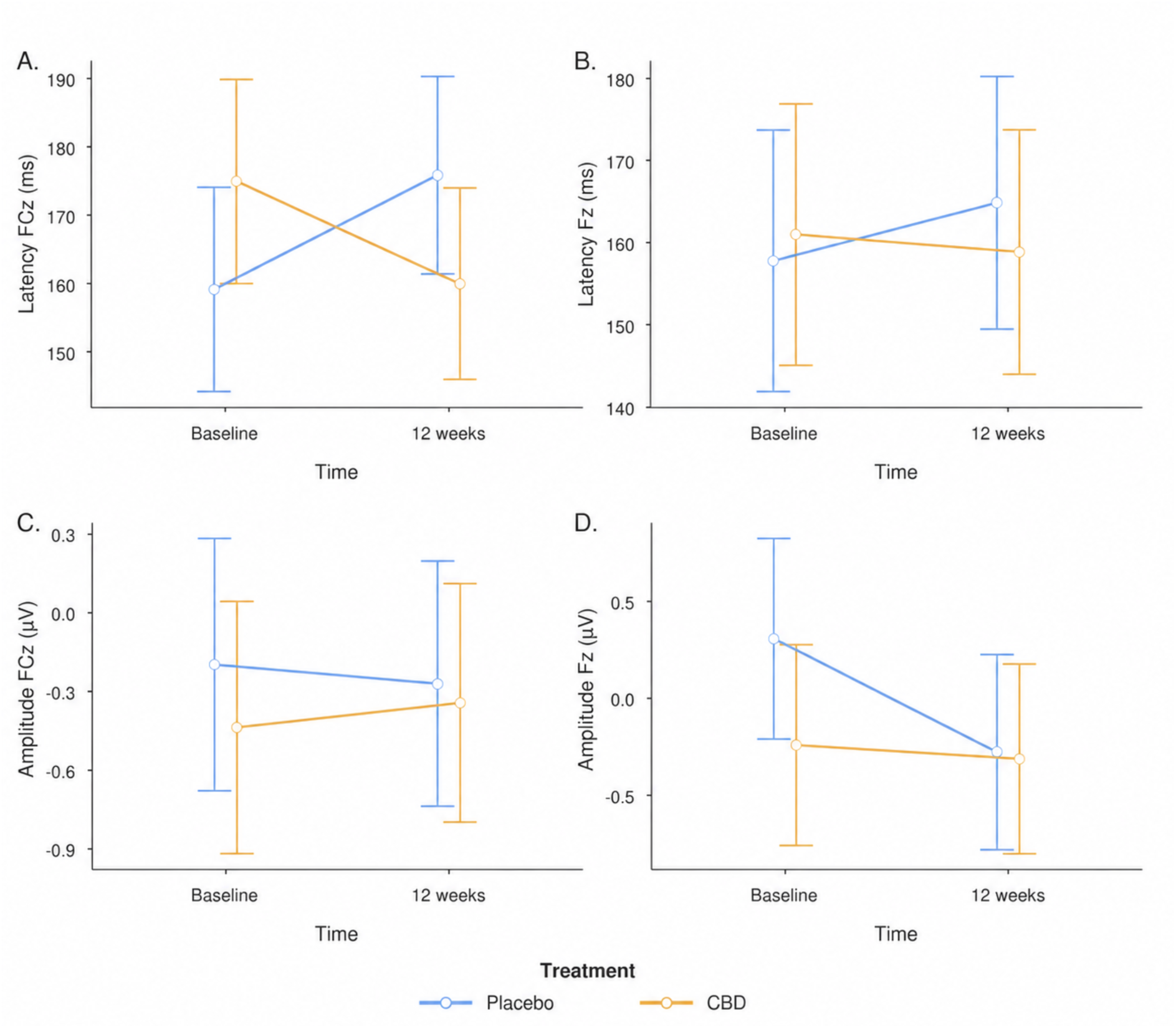
Estimated marginal means of MMN amplitude and peak latency across time and treatment at Fz and FCz. Points represent estimated marginal means from age-adjusted linear mixed-effects models; error bars represent 95% confidence intervals. CBD = cannabidiol; MMN = mismatch negativity.

**Table 2.** Estimated Marginal Means and Time × Treatment Effects for MMN Outcomes.

| Outcome | Electrode | CBD | | Placebo | | Time × Treatment | | | |
| --- | --- | --- | --- | --- | --- | --- | --- | --- | --- |
| | | Baseline | 12 weeks | Baseline | 12 weeks | Estimate [95% CI] | $F(df1, df2)$ | $p$ | $p_{FDR}$ |
| MMN latency (ms) | FCz | 174.9 | 160.0 | 159.1 | 175.8 | -31.7 [-59.8, -3.6] | 5.11(1, 40.12) | .029 | .058 |
| MMN latency (ms) | Fz | 161.0 | 158.8 | 157.8 | 164.8 | -9.2 [-40.3, 21.9] | 0.35(1, 54) | .556 | .556 |
| MMN amplitude ($\mu V$) | FCz | -0.44 | -0.34 | -0.20 | -0.27 | 0.17 [-0.66, 0.99] | 0.16(1, 38.4) | .687 | .687 |
| MMN amplitude ($\mu V$) | Fz | -0.24 | -0.31 | 0.31 | -0.28 | 0.51 [-0.37, 1.40] | 1.35(1, 40.73) | .251 | .502 |

*Note.* MMN = mismatch negativity; CBD = cannabidiol; FDR = false discovery rate; CI = confidence intervals. Estimated marginal means are adjusted for age. FDR correction was applied separately to amplitude and latency outcomes.

### MMN latency

At FCz, MMN peak latency decreased following CBD from 174.9 to 160.0 ms, whereas it increased from 159.1 to 175.8 ms following placebo (see Figure 3). The Time × Treatment interaction reached statistical significance, *F*(1, 40.12) = 5.11, *p* = .029, but did not survive FDR correction, *p*_FDR_ = .058. Given the apparent baseline difference between treatment conditions, we compared FCz MMN latency between the CBD and placebo conditions at baseline. The difference was not significant, *F*(1, 42.02) = 2.41, *p* = .128.

No significant Time × Treatment interaction was observed for MMN peak latency at Fz, *F*(1, 54.00) = 0.35, *p* = .556, *p*_FDR_ = .556. For this model, the participant random-intercept variance was estimated at zero (ICC = .00), indicating no detectable residual between-participant variability after accounting for the fixed effects. The random intercept was retained to reflect the repeated-measures structure of the data and maintain a consistent model specification across electrodes. Given the differing patterns at Fz and FCz, we conducted an exploratory follow-up model including Electrode (Fz, FCz) and its interactions with Time and Treatment. The Time × Treatment × Electrode interaction was not significant, *F*(1, 94.73) = 1.09, *p* = .299.

### MMN amplitude

No significant Time × Treatment interactions were observed for MMN mean amplitude at either FCz, *F*(1, 38.63) = 0.16, *p* = .687, *p*_FDR_ = .687, or Fz, *F*(1, 40.73) = 1.35, *p* = .251, *p*_FDR_ = .502.

## Exploratory Associations

Age-adjusted Spearman correlations at T0 showed that associations between DBC-2 Social Relating scores and MMN mean amplitude remained significant after FDR correction (see Table 3). Higher Social Relating scores were associated with more negative MMN amplitude at FCz, *ρ* = -.58, *p* = .028, *p*_FDR_ = .028, and Fz, *ρ* = -.59, *p* = .027, *p*_FDR_ = .028 (see Figure 4). Social Relating scores were not significantly associated with MMN peak latency at FCz, *ρ* = .30, *p* = .304, *p*_FDR_ = .304, or Fz, *ρ* = .44, *p* = .120, *p*_FDR_ = .240.

**Figure 4.**
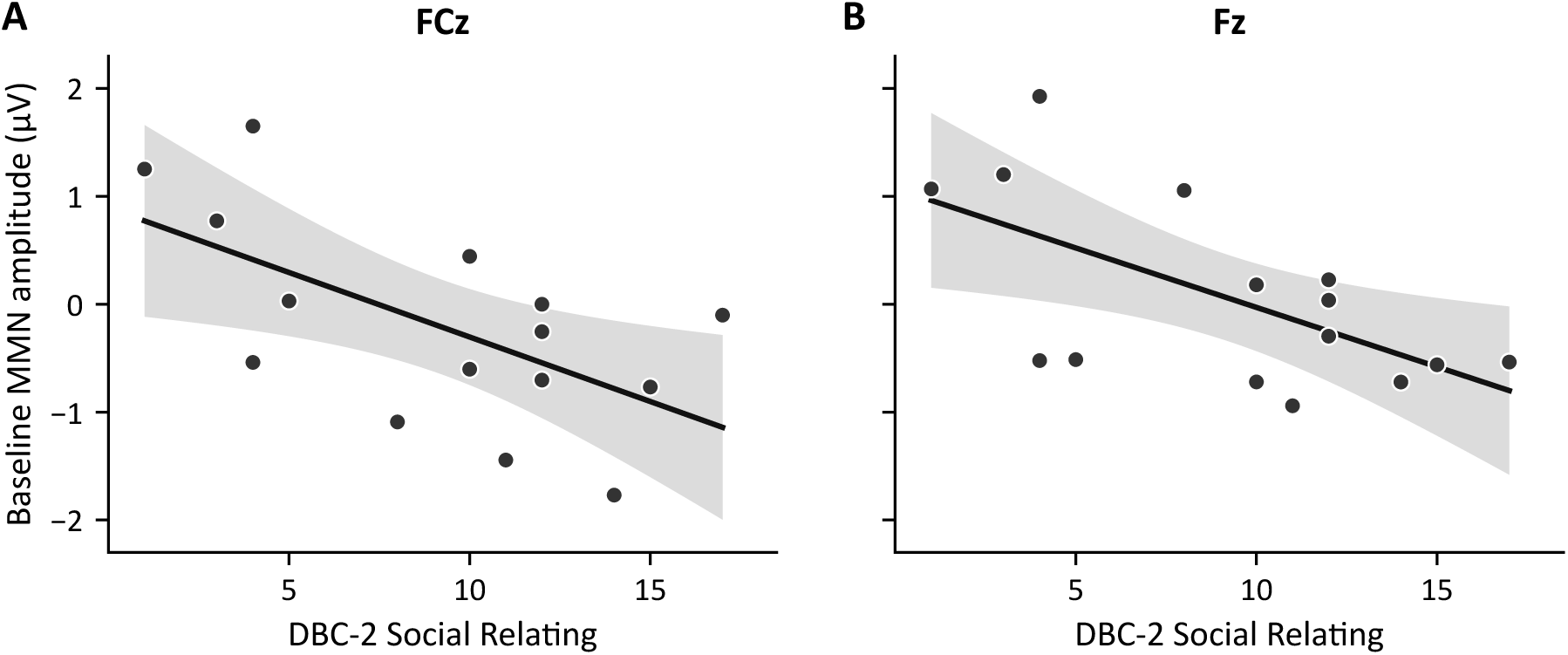
Associations between baseline MMN mean amplitude and DBC-2 Social Relating Scores at T0. Scatterplots show associations at (A) FCz and (B) Fz. Points represent individual participants’ raw values (*n* = 15). Solid lines show fitted linear regressions, with shaded regions indicating 95% confidence intervals. DBC-2 = Developmental Behaviour Checklist-Second Edition; MMN = mismatch negativity.

**Table 3.** Partial Spearman Correlations Between Baseline MMN Outcomes and DBC-2.

| Variable | 1 | 2 | 3 | 4 | 5 |
| --- | --- | --- | --- | --- | --- |
| 1. FCz latency | — |  |  |  |  |
| 2. Fz latency | .566 | — |  |  |  |
| 3. FCz amplitude | -.585 | -.317 | — |  |  |
| 4. Fz amplitude | -.516 | -.180 | .790 | — |  |
| 5. DBC-2 Social Relating (T0) | .296 | .435 | -.584* | -.588* | — |

*Note.* MMN = mismatch negativity; DBC-2 = Developmental Behaviour Checklist-Second Edition; T0 = initial baseline assessment. Values are partial Spearman's $\rho$ , controlling for age. False discovery rate (FDR) correction was applied separately within the amplitude and latency outcome families. \* $p_{\text{FDR}} < .05$ .

## Discussion

This study is among the first to examine whether multi-week CBD treatment modulates pre-attentive auditory deviance detection, indexed by MMN amplitude and latency, in autistic children. The study extends previous cannabinoid research reporting MMN modulation following acute administration in non-autistic adults (Greenwood et al., 2022; Juckel et al., 2007) by examining multi-week CBD treatment in a paediatric autistic sample. At FCz, the Time × Treatment interaction was consistent with the predicted reduction in MMN latency, but the effect did not survive correction for multiple comparisons and should therefore be interpreted cautiously. No corresponding effect was observed at Fz. The amplitude prediction was not supported, with no evidence of treatment-related differences in MMN amplitude at either electrode. Exploratory baseline analyses showed significant moderate-to-strong negative associations between social-relating scores and MMN amplitude at Fz and FCz that survived FDR correction, whereas associations with latency were nonsignificant.

Although the latency effect did not survive correction for multiple comparisons, its direction may reflect an earlier emergence of auditory mismatch processing following CBD treatment. Within a predictive-processing framework, one possibility is that CBD altered the postsynaptic gain of auditory prediction-error signals, thereby influencing the timing of precision-weighted processing (Brown & Friston, 2012; Lieder et al., 2013). Because postsynaptic gain depends partly on NMDAR-dependent recruitment of GABAergic circuitry and broader E/I control (Ferguson & Cardin, 2020; Lewis et al., 2022), this provides a potential link to CBD’s reported effects on glutamatergic and GABAergic signalling (Pretzsch et al., 2019; Zhang et al., 2023). This interpretation is broadly consistent with evidence that duration-deviant MMN latency is sensitive to NMDAR function (Kompus et al., 2015), with NMDAR antagonism generally associated with prolonged latency (Guo et al., 2024; Rosburg & Kreitschmann-Andermahr, 2016). Although CBD is not a direct NMDAR agonist, preclinical evidence suggests that repeated CBD treatment can attenuate behavioural and molecular disruptions induced by chronic NMDAR antagonism (Brakatselos et al., 2021; Gomes et al., 2015). Moreover, an altered glutamate-to-GABA ratio has also been associated with delayed MMN latency and greater interpersonal and communication difficulties (Ford et al., 2020). Together, these findings provide indirect support for an E/I-related interpretation, although the proposed mechanisms were not directly measured and remain speculative.

Previous cannabinoid studies have generally reported no significant differences in MMN latency. Such findings have been reported following acute administration of THC or THC-containing cannabis extract in healthy adults (Juckel et al., 2007) and in observational studies of chronic cannabis use (Roser et al., 2010). Conversely, delayed duration-deviant MMN latency has been reported in cannabis-using adults with early psychosis (Pesa et al., 2012). However, direct comparisons with the direction of the present finding are limited by differences in clinical context (e.g., reported alterations in endocannabinoid profiles in autism and psychosis; Karhson et al., 2018; Minichino et al., 2019) and age-related variation in MMN latency (Sapey-Triomphe et al., 2025). Cannabinoid composition, dosing regimen, and prior exposure may also be relevant. Unlike previous studies involving acute experimental administration of THC-containing preparations or recreational cannabis use, the present study examined repeated therapeutic CBD treatment in cannabinoid-naive autistic children. This distinction may be important because CBD and THC have shown differing effects on brain function. For instance, CBD has been reported to attenuate some THC-related effects (Gunasekera et al., 2021) and has established antiseizure efficacy, whereas THC has shown some proconvulsant effects (Devinsky et al., 2024; Kaczor et al., 2022). Prior cannabis exposure may also influence MMN responses, as CBD-related MMN effects have been observed selectively in adults with less frequent cannabis use (Greenwood et al., 2022).

The latency reduction appeared more pronounced at FCz than Fz, although the treatment effect did not differ statistically between electrodes. One possibility is that CBD altered the relative timing of activity within the frontotemporal MMN network, in which auditory cortical activity generally precedes later frontal contributions (Fitzgerald & Todd, 2020). Auditory-cortical contributions appear particularly prominent in childhood, with greater temporal MMN activity in school-aged children than in adults (Gomot et al., 2000) and MMN responses localised to auditory cortex in children (Ruhnau et al., 2013). Because CBD modulates temporal cortical activity during auditory processing (Ghosh et al., 2021; Winton-Brown et al., 2011), altered timing of this activity could shift the scalp-recorded MMN differently at FCz and Fz, as each reflects a different weighting of underlying neural sources (Michel & Brunet, 2019). However, given the non-significant electrode comparison, the apparent FCz-specific pattern may instead reflect sampling variability, or our modest sample may have limited sensitivity to such a difference. Without a larger sample and source-level measures, we cannot distinguish between these possibilities.

The absence of an amplitude effect may reflect the context-sensitive nature of the MMN. Because MMN responses in autism vary across paradigms and deviant type (Cacciato-Salcedo et al., 2026), CBD-related modulation may also depend on stimulus characteristics. Notably, the duration deviant may have imposed different processing demands on the auditory system (Takasago et al., 2024). Whereas frequency and intensity deviants can be detected from stimulus onset, a longer-duration deviant can only be identified once the tone extends beyond its expected offset. Duration processing may therefore rely more on tracking elapsed time against an internal temporal expectation than on an immediate acoustic comparison (Moberget et al., 2008; Takasago et al., 2024). This may have made CBD-related effects more apparent in latency than amplitude. This interpretation is broadly consistent with evidence that NMDAR antagonism can alter duration-MMN latency without a corresponding amplitude effect, whereas amplitude effects are more consistently observed for frequency and intensity deviants (Guo et al., 2024). Given our study’s limited sensitivity to smaller amplitude interactions, larger future studies incorporating multiple deviant types could test whether CBD-related MMN modulation differs across duration, frequency, and intensity changes.

Exploratory baseline analyses showed that higher DBC-2 Social Relating scores, reflecting greater social-relating difficulty, were associated with more negative MMN amplitude at both Fz and FCz, with not with latency. This is consistent with evidence that relationships between MMN measures and autistic characteristics vary in direction (Sapey-Triomphe et al., 2025). Both larger and smaller MMN amplitudes have been associated with greater autistic traits or atypical sensory features (Ludlow et al., 2014; Matsuba et al., 2024), and both shorter and longer latencies with language impairment or communication difficulties (Green et al., 2020; Matsuzaki et al., 2019). Although precision was not measured directly, the amplitude relationship may reflect individual differences in the precision assigned to duration-based auditory prediction errors across variation in social-relating difficulty (Cannon et al., 2021). More broadly, these findings caution against interpreting larger MMN amplitude or shorter latency as inherently reflecting more or less adaptive social and behavioural functioning in autism.

## Limitations and future research

Several limitations should be considered when interpreting the present findings. First, because EEG was a secondary outcome of the broader trial, relatively few participants contributed usable EEG data. The sensitivity analysis indicated limited power to detect smaller treatment-related effects, and the limited number of paired observations precluded examining whether CBD-related MMN changes covaried with changes in social-relating difficulties. Adequately powered trials could test whether CBD reliably modulates MMN and whether such modulation tracks meaningful changes in social functioning, clarifying its potential as a neurophysiological marker of treatment response.

Second, MMN was examined using a single duration deviant, so it remains unclear whether the latency effect without a corresponding amplitude change is specific to duration processing. Future trials incorporating multiple deviant types could determine whether CBD-related MMN modulation generalises across auditory features. Such work could identify which MMN parameters and deviant types are most sensitive to CBD and therefore most suitable as outcome measures in future trials.

Third, the proposed neurobiological mechanisms and scalp-recorded MMN cannot establish whether CBD altered glutamatergic and GABAergic signalling or activity within the frontotemporal network. Combining repeated source-localised EEG with magnetic resonance spectroscopy measures of auditory cortical glutamate and GABA could test whether CBD-related MMN modulation covaries with neurochemical changes and with altered timing or contributions of temporal and frontal generators. Follow-up assessments beyond the 12-week intervention could further determine whether the changes persist after treatment and are accompanied by behavioural improvement, clarifying the durability and clinical relevance of CBD-related effects.

## Conclusion

Overall, this secondary analysis of EEG data found tentative evidence that 12 weeks of CBD treatment may modulate automatic auditory deviance detection in autistic children, with the observed pattern confined to duration-MMN latency rather than amplitude. Exploratory analyses also showed that greater baseline social-relating difficulties were associated with more negative MMN amplitude, suggesting that individual differences in auditory deviance processing may relate to social functioning in autism. Although the treatment-related latency finding did not survive correction for multiple comparison, this study extends previous cannabinoid research by examining MMN following repeated CBD administration in autistic children and highlights the potential value of incorporating objective neurophysiological measures alongside caregiver- and clinician-reported outcomes in future CBD trials.

## Data Availability

All data produced in the present study are available upon reasonable request to the authors

## Acknowledgement

The investigational product was provided in-kind by Medigrowth Australia.

