## Supplementary material for "Effects of Cannabidiol on Auditory Mismatch Negativity in Autistic Children"

**Supplementary material: Effects of Cannabidiol on Auditory Mismatch Negativity in Autistic Children**

**Figure 1**

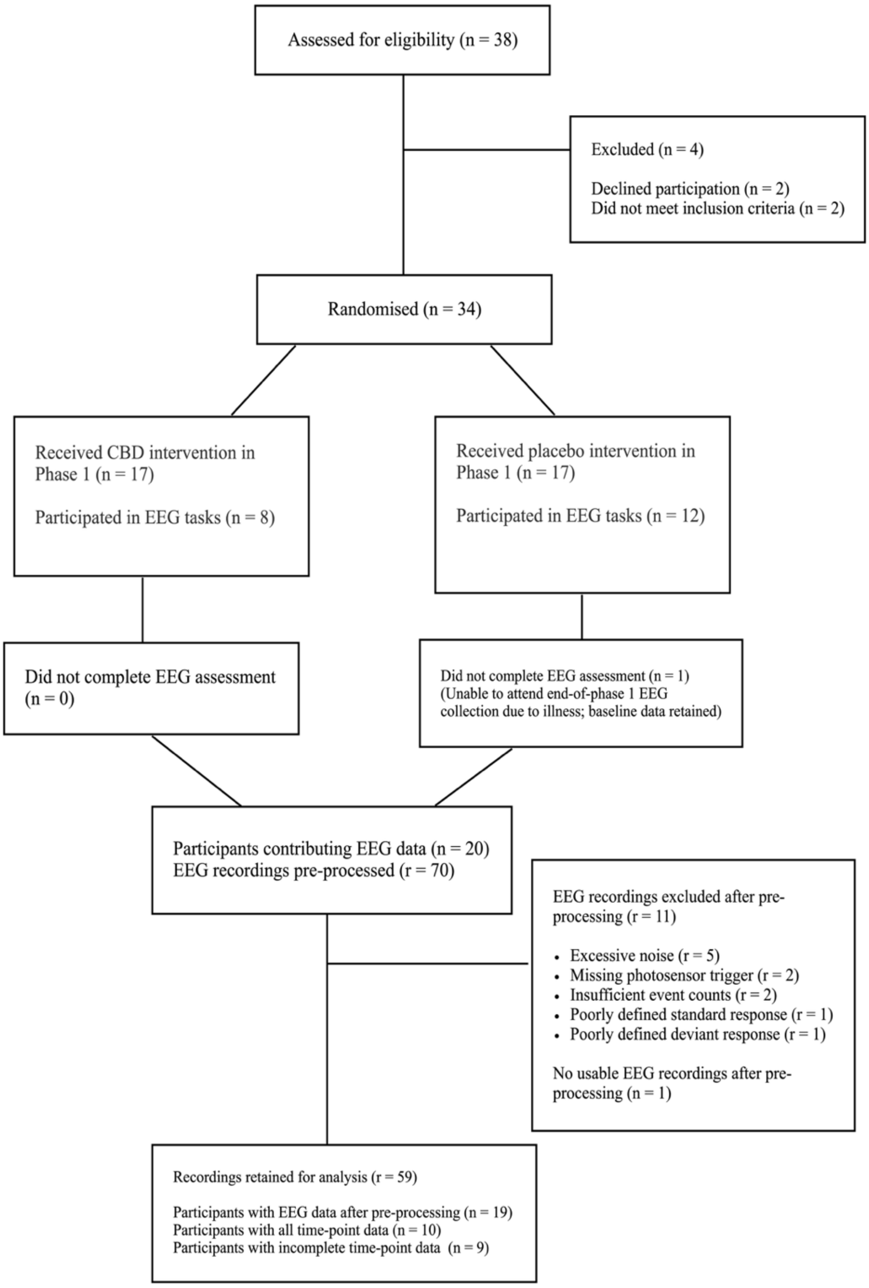


*CONSORT diagram depicting participant flow and EEG data retention.* *r* = number of recordings; *n* = number of participants; EEG = electroencephalography; CBD = cannabidiol.
